# Tenecteplase 0.4 mg/kg in moderate and severe acute ischemic stroke: A pooled analysis of NOR-TEST & NOR-TEST 2A

**DOI:** 10.1101/2023.05.04.23289542

**Authors:** Vojtech Novotny, Christopher Elnan Kvistad, Halvor Naess, Nicola Logallo, Annette Fromm, Andrej Netland Khanevski, Lars Thomassen

**Author notes:** **Corresponding author:** Vojtech Novotny, **E-mail:**, **Address:** Department of Neurology, Haukeland University Hospital; Jonas Lies vei 65; N-5021 Bergen, Norway, **Twitter:** @vojtech_no.

## Abstract

**Background:** The optimal dose of tenecteplase in acute ischaemic stroke remains to be defined. We present a pooled analysis of the two NOR-TEST trials exploring the efficacy and safety of tenecteplase 0.4 mg/kg in acute ischemic stroke.

**Methods:** We retrospectively reviewed two multi-center PROBE trials, NOR-TEST and NOR-TEST 2A, conducted in Norway. The patients were randomized to either 0.4 mg/kg single bolus tenecteplase or standard 0.9 mg/kg alteplase. The primary endpoint was favorable functional outcome at three months (mRS 0-1) or return to baseline if pre-stroke mRS was 2. The secondary endpoints included favorable functional outcome at three months (Modified Rankin Scale 0-2), major neurological improvement and safety data. The pooling project includes a pooled analysis of patients with moderate to severe stroke (NIHSS ≥6) from both trials and an additional post-hoc analysis of patients with mild stroke (NIHSS ≤5) from NOR-TEST.

**Results:** The per-protocol analysis contains 483 patients, of which 235 were assigned to tenecteplase and 248 to alteplase. In per-protocol analysis, functional outcome was better in the alteplase arm with cut-off mRS 2 (OR 0.52, 95% CI 0.33-0.80, p=0.003) and expressed by ordinal shift analysis (OR 1.64, 95% CI 1.17-2.28, p=0.004). Mortality at 3 months was higher in the tenecteplase arm (OR 2.48, 95% CI 1.20-5.10, p=0.014). Mortality and intracranial hemorrhage rates were higher in the severe stroke group randomized to tenecteplase, whereas these rates were similar for alteplase and tenecteplase in moderate and mild stroke. High age was not associated with either higher mortality or intracranial hemorrhage rates.

**Conclusions:** Tenecteplase 0.4 mg/kg is unsafe in moderate and severe stroke and the risk of death and intracranial hemorrhage probably increases with stroke severity. A lower tenecteplase dose should be tested in future trials.

**Trial Registration:** ClinicalTrials.gov Identifiers: NCT01949948, NCT03854500 https://clinicaltrials.gov/ct2/show/NCT01949948 https://clinicaltrials.gov/ct2/show/NCT03854500

## Introduction

Alteplase is beneficial in patients with acute ischemic stroke (AIS) in the whole age specter.^1, 2^ Tenecteplase as a thrombolytic agent is preferable over alteplase in several aspects.^3, 4^ Besides better biochemical features, including longer half-life and higher fibrin specificity, the single bolus administration makes tenecteplase an attractive thrombolytic in the acute setting, especially with the “drip and ship” strategy for patients with large vessel occlusions in need of transport for mechanical thrombectomy.

In recent years, there have been increasing research activities aiming to replace alteplase with tenecteplase for AIS. Several phase 2 trials comparing tenecteplase with alteplase, with varying inclusion criteria and tenecteplase doses, have been performed. They have in most cases shown similar efficacy and safety profile of the two thrombolytics. The Norwegian tenecteplase trials NOR-TEST and NOR-TEST 2A tested tenecteplase 0.4 mg/kg since data on the ideal dose were inconclusive at that time and based on small sample sizes or highly selected patients in previous studies. ^5-11^ Accumulated clinical trials indicate that tenecteplase may be preferable compared with alteplase in the treatment of AIS.^12^ However, the ideal dose has become a key question. NOR-TEST was the first phase 3 trial comparing tenecteplase 0.4 mg/kg with standard alteplase 0.9 mg/kg.^13^ However, the cohort contained an unproportionally high number of patients with mild AIS and stroke mimics. The results were therefor difficult to apply to the general stroke population. However, the data encouraged continued research.^14^ A sub-analysis of patients with moderate and severe AIS in NOR-TEST showed similar rates of favorable outcome and symptomatic intracranial hemorrhage (sICH) in both treatment arms, although the mortality rate at 3 months was higher in severe AIS.^15^ Based on these data, closer monitoring of safety parameters in subsequent trials testing tenecteplase 0.4 mg/kg was recommended. NOR-TEST 2A was designed to clarify non-inferiority of tenecteplase in patients with moderate and severe AIS using the 0.4 mg/kg dose. The trial was, however, prematurely terminated when a per-protocol safety analysis of the first 200 patients showed worse safety and functional outcomes in patients treated with tenecteplase 0.4 mg/kg compared with those treated with alteplase 0.9mg/kg.^16^ We therefore performed a pooled analysis of both NOR-TEST trials, in order to illuminate efficacy and safety of tenecteplase 0.4 mg/kg based on a larger cohort of patients.

## Methods

### Design of the RTCs and Subjects

The Norwegian Tenecteplase Stroke Trials “NOR-TEST” (NCT01949948) & “NOR-TEST 2A” (NCT03854500) were multicenter, phase 3, randomized, open-label, blinded endpoint trials.^13, 16^ NOR-TEST was performed at 13 sites and NOR-TEST 2A at 11 sites. Patients were enrolled into NOR-TEST from September 1, 2012, until September 30, 2016, and in NORTEST 2A from October 28, 2019, until September 26, 2021.

The two trials contain altogether 1323 patients of which 1107 patients from NOR-TEST (83.7%) and 216 patients from NOR-TEST 2A (16.3%). For the intention-to-treat (ITT) analysis, 19 patients were excluded either because of withdrawal of informed consent after inclusion, or due to reconsideration of eligibility prior to medication administration, resulting in a final number of 1304 patients (1100 from NOR-TEST and 204 from NOR-TEST 2). The randomization allocated 649 patients to tenecteplase (49.8%) and 655 patients to alteplase (50.2%). The ITT represents all the subjects included in the trials regardless of their final diagnosis. The per-protocol (PP) analysis excludes all patients not matching the inclusion criteria, i.e. patients with other diagnosis than ischemic stroke, modified Rankin Scale ≥3, NIHSS <6, and patients missing primary outcome data for the final analysis.

The inclusion criteria in both trials differed in terms of National Institutes of Health Stroke Scale (NIHSS) score on admission. NOR-TEST included all eligible patients with suspected AIS and a neurological deficit measurable by NIHSS, whereas NOR-TEST 2A included only patients with NIHSS ≥6 on admission. Otherwise, the inclusion criteria were identical. Patients ≥18 years of age, with modified Rankin Scale (mRS) 0-2 prior the admission and who were admitted within 4.5 hours from stroke onset were eligible for study inclusion. Patients with wake-up stroke or unknown time of stroke onset were considered eligible for study inclusion when Diffusion Weighted Imaging (DWI) - Fluid Attenuated Inversion Recovery (FLAIR) mismatch was proven on admission Magnetic Resonance Imaging (MRI). Patients receiving bridging thrombolytic therapy prior endovascular treatment were eligible for inclusion. Patients were randomized 1:1 to either 0.4 mg/kg single bolus tenecteplase (maximum dose of 40 mg) or to standard alteplase dose 0.9 mg/kg (10% bolus and 90% infusion over 60 minutes with maximum dose of 90 mg). The treating staff in the emergency room was not blinded to treatment randomisation, but health personnel in the stroke unit and at follow-up were. Patients were unaware which drug they had received. Further details regarding randomization and procedures were published previously.^16, 17^

The pooling project includes a pooled analysis of patients with moderate (NIHSS 6–14) or severe (NIHSS ≥15) AIS from NOR-TEST and NOR-TEST 2A, and an additional post-hoc analysis of patients with mild AIS (NIHSS ≤5) from NOR-TEST.

### Outcomes

The primary endpoint was favorable functional outcome at 3 months defined as mRS 0-1 or return to baseline if pre-stroke mRS was 2. The secondary endpoints were favorable functional outcome at 3 months defined as mRS 0-2; major neurological improvement at 24 hours measured by NIHSS; any intracranial hemorrhage (ICH) and sICH occurring within 24-48 hours after symptom onset; (favorable) ordinal shift analysis of mRS at 3 months; and mortality within 3 months. Any ICH was defined as any hemorrhagic transformations or parenchymal hematoma according to European Cooperative Acute Stroke Study (ECASS) I criteria.^18^ ICH morphology was described according to ECASS I criteria.^19^ SICH was defined according to ECASS III criteria.^20^ Major neurological improvement was defined as either NIHSS score of 0 at 24 h or a reduction in NIHSS score of at least 4 points at 24 hours compared with baseline. In this post-hoc analysis, we stratified the cohort into three age groups, i.e. ≤60, 60-80, ≥80 years, and three stroke severity groups, i.e. mild, moderate and severe.

### Statistics

Primary and secondary endpoints were essentially identical for both trials and were analyzed as ITT and PP analysis adjusted for age, pretreatment NIHSS, premorbid mRS, time from onset to IVT, endovascular treatment and source trial. However, the PP analysis is presented in the graphics as the main result in this study. For the demographics, the continuous variables were tested by t-test in case of normally distributed data and by Mann–Whitney U-test in case of uneven distribution. Variations were expressed by standard deviation or interquartile range, respectively. The categorical variables were tested by Pearson’s chi-squared test. For the final analyses, a logistic regression analysis expressed by odd ratio (OR) was used. The significance of p-value was set to < 0.05. The primary and secondary outcomes are illustrated using appropriate histograms. An additional post-hoc analysis of patients with mild stroke from NOR-TEST was included in the graphics to better illustrate an overall difference between the stroke severity groups. Anonymized data supporting our findings in the presented article may be provided by Vojtech Novotny or Christopher Elnan Kvistad upon reasonable request.

The study was approved by the regional Committee for Medical and Health Research Ethics and the Norwegian Medicines Agency. Written informed consent was obtained from study participants or their legal representatives in both trials. The funding company had no role in study design, data analysis, interpretation or writing of the manuscript.

## Results

In the pooled analysis of patients with moderate or severe stroke, the ITT population contained 597 patients of which 287 (48.1%) were assigned to tenecteplase and 310 (51.9%) to alteplase. The exclusion scheme for the PP analysis followed the criteria of the original trials. The following patients were excluded from the ITT population: 60 (10%) patients diagnosed as stroke mimic, 36 (6%) patients with pre-stroke mRS ≥ 3, 17 (3%) patients with missing end-point data at 3 months, and one patient having NIHSS < 6. The final PP population included 483 patients, of which 235 (48.7%) were assigned to tenecteplase and 248 (51.3%) to alteplase (Figure S1). The demographics between the two treatment arms were similar, except for a higher occurrence of pre-stroke myocardial infarction in the alteplase arm (Table 1).

**Table 1.** Demographics and characteristics in the per-protocol analysis

|  | **Tenecteplase N=235** | **Alteplase N=248** | **P value** |
| --- | --- | --- | --- |
| **Age (years)** |  |  |  |
| Mean (SD) | 70.8 (13.9) | 70.5 (13.8) | 0.817 |
| Median (IQR) | 73 (62-81) | 73 (62-80) | 0.849 |
| **Weight (Kg)** |  |  |  |
| Mean (SD) | 77.8 (15.2) | 78.95 (14.5) | 0.519 |
| Median (IQR) | 76 (68-87) | 80 (70-88.5) | 0.573 |
| **Age groups (years)** |  |  |  |
| <60, N (%) | 52 (22.1) | 53 (21.4) | 0.840 |
| 60-80, N (%) | 122 (51.9) | 134 (54.0) | 0.641 |
| >80 (N, %) | 61 (26.0) | 61 (24.6) | 0.731 |
| **Sex** |  |  |  |
| Women, N (%) | 105 (44.7) | 105 (42.3) | 0.604 |
| Men, N (%) | 130 (55.3) | 143 (57.7) |  |
| **Unknown time of stroke symptom onset, N (%)** | 19 (8.1) | 13 (5.3) | 0.209 |
| **Major arterial vessel occlusion, N (%)** | 97 (41.3) | 113 (45.6) | 0.342 |
| **Endovascular treatment, N (%)** | 45 (19.2) | 59 (23.8) | 0.215 |
| **Final diagnosis** |  |  |  |
| Ischemic stroke, N (%) | 220 (93.6) | 233 (93.95) | 0.879 |
| Transitory ischaemic attack, N (%) | 14 (5.96) | 13 (5.24) | 0.732 |
| Stroke mimics, N (%) | 0 (0) | 0 (0) | - |
| **Stroke risk factors** |  |  |  |
| Hypertension, N (%) | 120 (51.1) | 128 (51.6) | 0.904 |
| Atrial fibrillation, N (%) | 27 (11.5) | 36 (14.5) | 0.324 |
| Diabetes mellitus, N (%) | 29 (12.3) | 31 (12.5) | 0.958 |
| Hypercholesterolemia, N (%) | 49 (20.9) | 50 (20.2) | 0.851 |
| Smoker, N (%) | 54 (22.98) | 57 (22.98) | 0.999 |
| **Cardiovascular history** |  |  |  |
| Prior ischemic stroke, N (%) | 37 (15.7) | 35 (14.1) | 0.615 |
| Prior myocardial infarction, N (%) | 23 (9.8) | 42 (16.94) | 0.021 |
| **Premorbid mRS score** |  |  |  |
| 0, N (%) | 176 (74.9) | 196 (79.03) | 0.280 |
| 1, N (%) | 41 (17.45) | 36 (14.52) | 0.379 |
| 2, N (%) | 18 (7.66) | 16 (7.1) | 0.604 |
| ≥3, N (%) | - | - | - |
| **NIHSS score on admission** |  |  |  |
| Mean (SD) | 12.1 (6.5) | 12.1 (5.8) | 0.910 |
| Median (IQR) | 10 (7-15) | 10 (8-15) | 0.580 |
| Moderate (6-14) | 169 (71.91) | 176 (70.97) | 0.818 |
| Severe (≥15) | 66 (28.09) | 72 (29.03) | 0.818 |
| **TOAST classification** |  |  |  |
| Large vessel disease, N (%) | 69 (29.4) | 59 (23.8) | 0.166 |
| Cardioembolism, N (%) | 69 (29.4) | 83 (33.5) | 0.331 |
| Small vessel disease, N (%) | 19 (8.1) | 16 (6.5) | 0.489 |
| Other causes, N (%) | 12 (5.1) | 16 (6.5) | 0.527 |
| Unknown or several causes, N (%) | 63 (26.8) | 66 (26.6) | 0.961 |
| **Time (minutes) from onset to thrombolysis (IQR)** | 100 (70-146) | 94 (70-136) | 0.556 |

NIHSS: National Institutes of Health Stroke Scale; mRS: modified Rankin scale; SD:
Standard deviation; IQR: interquartile range; BP: blood pressure; TOAST: trial of ORG
10172 in acute stroke treatment

In the pooled population with moderate and severe stroke, the ITT analysis showed a trend towards higher rates of any ICH and sICH in patients treated with tenecteplase [OR 1.53, 95% CI 0.99 – 2.66, p=0.051) and (OR 2.51, 95% CI 0.98-6.44, p=0.054) respectively]. In the alteplase arm, ordinal shift analysis of mRS at 3 months showed better functional outcome (OR 1.18, 95% CI 1.18 – 2.18, p=0.002) as well as better functional outcome at 3 months using mRS cut-off 0-2 (OR 0.50, 95% CI 0.33 – 0.78, p=0.002). Major neurological improvement at 24 h expressed by NIHSS was more common in the alteplase arm (OR 0.77, 95% CI 0.49 – 0.99, p=0.05). Mortality at 3 months was more common in the tenecteplase arm (OR 2.42, 95% CI 1.28 – 4.59, p=0.007) (Table 2).

**Table 2.** Study outcomes and intracranial hemorrhage characteristics in the intention to treat analysis

|  | Intention-to-treat |  |  |  |
| --- | --- | --- | --- | --- |
|  | Tenecteplase<br>n=287 | Alteplase<br>n=310 | OR (95% CI) | P value |
| **Primary endpoint** |  |  |  |  |
| mRS score 0-1 at 3 months, N (%) | 118 (42.9) | 138 (45.9) | 0.78 (0.53 – 1.15) | 0.214 |
| **Secondary endpoints** |  |  |  |  |
| mRS score 0-2 at 3 months*, N (%) | 151 (54.9) | 192 (63.8) | 0.50 (0.33 – 0.78) | 0.002 |
| Major neurological improvement at 24 h expressed by NIHSS, N (%) | 162 (59.1) | 196 (65.3) | 0.77 (0.49 – 0.99) | 0.050 |
| Ordinal shift analysis of mRS at 3 months | - | - | 1.18 (1.18 – 2.18) | 0.002 |
| Any ICH, N (%) | 48 (16.7) | 36 (11.6) | 1.53 (0.99 – 2.66) | 0.051 |
| HI1, N (%) | 10 (3.5) | 6 (1.9) |  | 0.242 |
| HI2, N (%) | 11 (3.8) | 9 (2.9) |  | 0.528 |
| PH1, N (%) | 6 (2.1) | 8 (2.6) |  | 0.693 |
| PH2, N (%) | 9 (3.1) | 7 (2.3) |  | 0.507 |
| PHr, N (%) | 9 (3.1) | 4 (1.3) |  | 0.162 |
| IVH, N (%) | 1 (0.4) | 0 |  | 0.481 |
| Symptomatic ICH, N (%) | 15 (5.2) | 7 (2.3) | 2.51 (0.98 - 6.44) | 0.054 |
| mRS score 5-6 at 3 months, N (%) | 39 (14.2) | 30 (10.0) | 1.49 (0.99 – 3.07) | 0.055 |
| Mortality at 3 months, N (%) | 35 (12.7) | 21 (7.0) | 2.42 (1.28 – 4.59) | 0.007 |

ICH: intracranial hemorrhage; mRS: modified Rankin Scale; NIHSS: National Institutes of Health Stroke Scale; HI: Hemorrhagic infarction; PH: Parenchymal haemorrhage; PHr: Remote parenchymal hemorrhage; IVH: Intraventricular hemorrhage

The PP analysis showed in the alteplase arm a better functional outcome at 3 months expressed by ordinal shift analysis (OR 1.64, 95% CI 1.17 – 2.28, p=0.004) and a better functional outcome at 3 months using mRS cut-off 0-2 (OR 0.52, 95% CI 0.33 – 0.80, p=0.003). There was a higher rate of any ICH (OR 1.66, 95% CI 0.97 – 2.82, p=0.062) and sICH (OR 2.39, 95% CI 0.79 – 7.24, p=0.121) in the tenecteplase arm, but none of these reached statistical significance. Mortality was significantly higher in the tenecteplase arm (OR 2.48, 95% CI 1.20 – 5.10, p=0.014) (Table 3 & Figure 1).

**Table 3.**
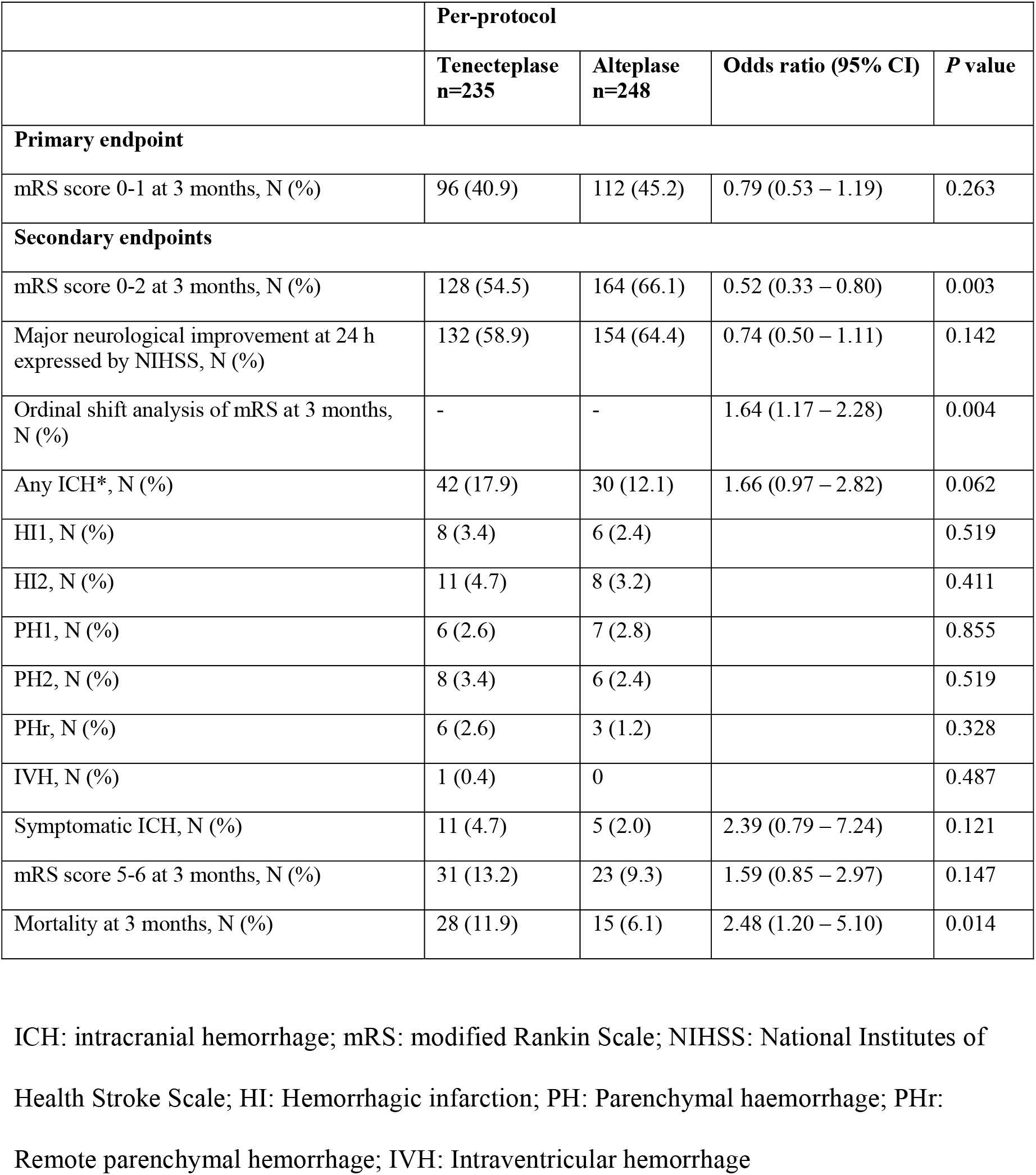
Study outcomes and intracranial hemorrhage characteristics in the per-protocol analysis

**Figure 1.**
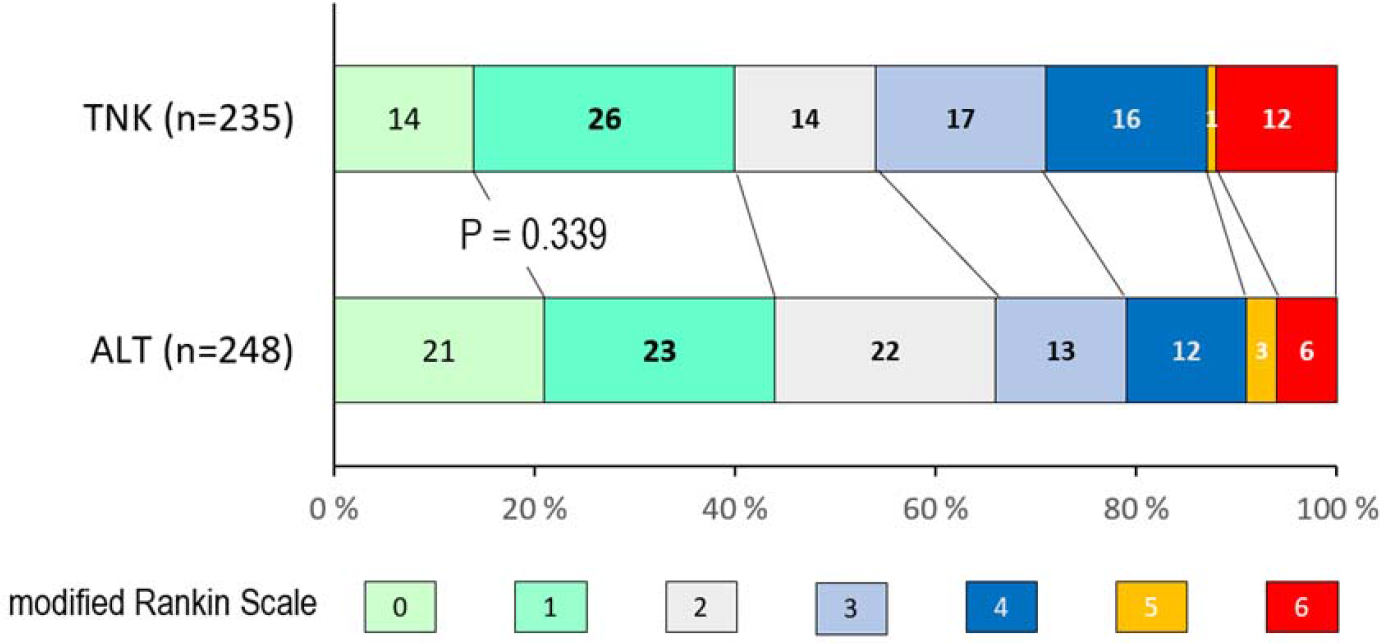
Distribution of modified Rankin Scale scores at 3 months in the per protocol analysis TNK: tenecteplase; ALT: alteplase

When stratified by age, patients 60 – 80 years of age in the tenecteplase arm showed more any ICH and sICH (22.1% vs. 9% and 8.2% vs. 1.5%, respectively) and a higher mortality rate (9.8% vs. 3.7%), whereas favorable functional outcome was more common in the alteplase arm (56.1% vs. 40.2%). There was no difference regarding ICH rates or functional outcome in the age groups <60 years and >80 years (Figure 2).

**Figure 2.**
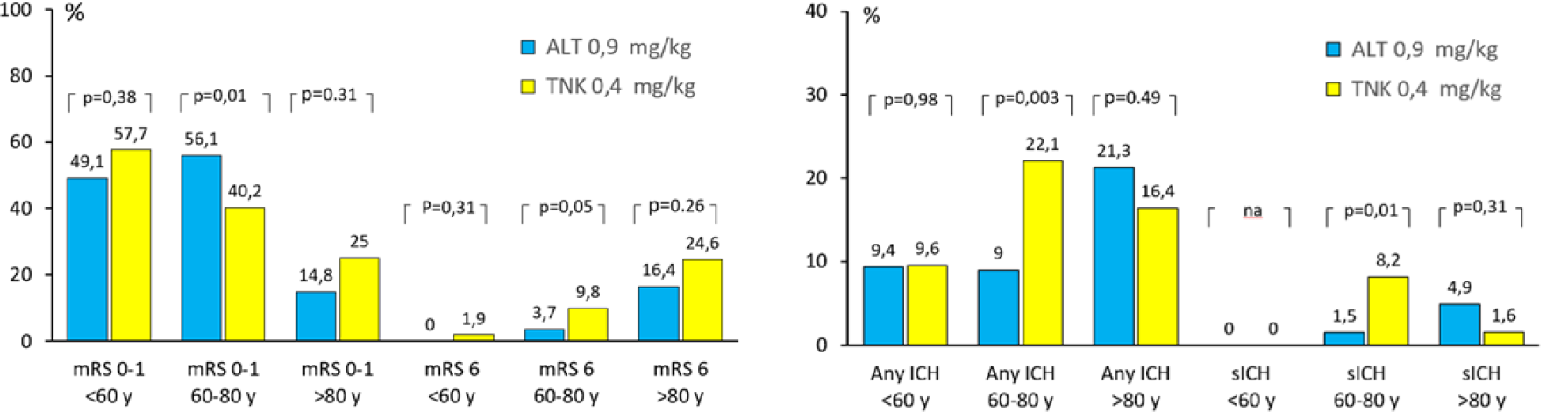
Distribution of outcome and hemorrhage based on age groups in the per protocol analysis mRS: modified Rankin Scale; ALT: alteplase; TNK: tenecteplase; y: years; ICH: intracranial hemorrhage; sICH: symptomatic intracranial hemorrhage.

When stratified by stroke severity, favorable outcome was similar in both treatment arms, but mortality in patients with severe stroke was higher in the tenecteplase arm (27.3% vs. 8.3%). Both any ICH (31.8% vs. 15.3%) and sICH (9.1% vs. 1.4%) were more common in patients with severe stroke treated with tenecteplase. Patients with mild and moderate stroke had similar rates of any ICH and sICH regardless of the type of thrombolytic treatment (Figure 3).

**Figure 3.**
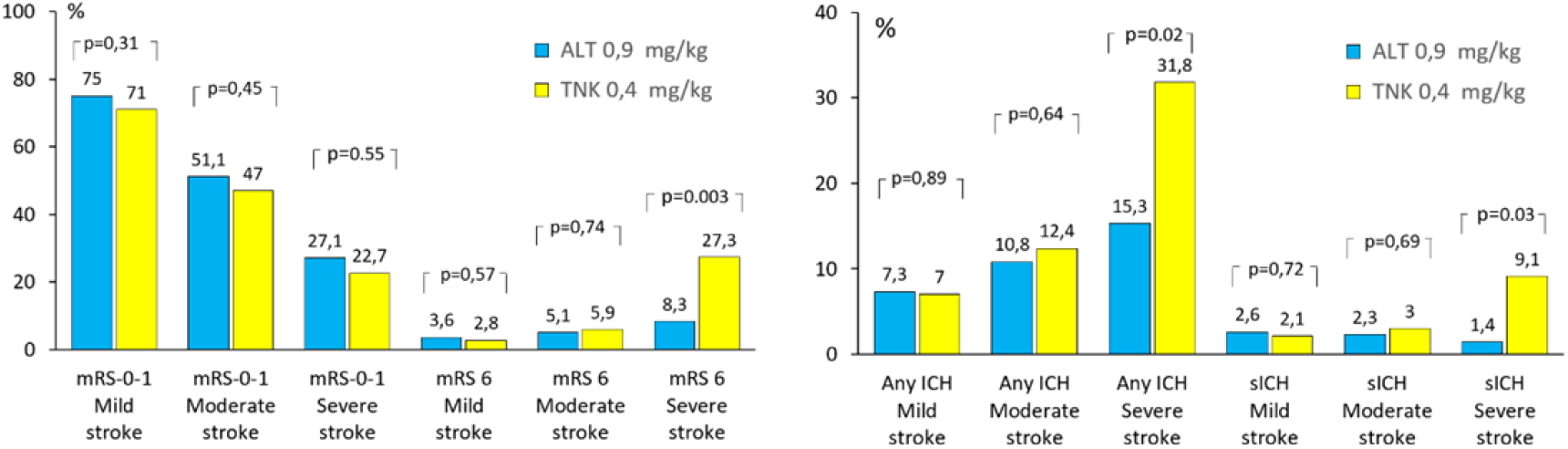
Distribution of outcome and hemorrhage based on severity of stroke on admission in the per protocol analysis mRS: modified Rankin Scale; ALT: alteplase; TNK: tenecteplase; Mild stroke: NIHSS≤5; Moderate stroke: NIHSS 6–14; Severe stroke: NIHSS ≥15; ICH: intracranial hemorrhage; sICH: symptomatic intracranial hemorrhage; NIHSS: National Institutes of Health Stroke Scale

Compared to the main analysis, there was no significant change in the primary or secondary outcomes after exclusion of patients with unknown stroke onset (Table S1). In the separate analysis on patients undergoing endovascular treatment, patients treated with alteplase achieved more often major neurological improvement (OR 0.34, 95% CI 0.12 – 0.93, p=0.035) and any ICH occurred less often (OR 1.70, 95% CI 1.7 – 14.65, p=0.003) (Table S2).

In the tenecteplase arm, ITT analysis showed that death within 90 days occurred in 35 patients. Among 22 patients with severe stroke, 12 patients died of the initially large ischemic stroke and malignant edema; two of sICH; one of renal insufficiency and pneumonia; one of herpes encephalitis; and 6 of unknown cause in nursing homes. Among 13 patients with moderate stroke, two patients died of sICH; one of lung embolism; one of myocardial infarction; one of cardiac failure, one of fatal recurrent major ischemic stroke; and 7 of unknown cause in nursing homes.

In the alteplase arm, death within 90 days occurred in 21 patients. Among 9 patients with severe stroke, two patients died of sICH; four of the initially large ischemic stroke and malignant edema; one of pneumonia; and two of unknown cause in nursing homes. Among 12 patients with moderate stroke, three patients died of sICH; one of myocardial infarction and cardiac failure; one of pneumonia; and seven of unknown cause in nursing homes.

## Discussion

The pooled data analysis of both NOR-TEST trials confirms the conclusion of NOR-TEST 2A, stating that tenecteplase 0.4 mg/kg is not safe in patients with moderate and severe AIS. The pooled analysis was performed mainly to reduce potential bias. The pooled analysis contains, in contrast to NOR-TEST, almost 50% fewer stroke mimics.^13^ Furthermore, the primary and secondary endpoints in the pooled analysis were analyzed, as in NOR-TEST 2A, excluding mild stroke patients.^16^ Both mild stroke patients and stroke mimics have a significantly better prognosis and lower occurrence of sICH compared to moderate and severe stroke which statistically obscures the true safety profile of the tested thrombolytics.^21, 22^

NOR-TEST 2A was designed with a firm power calculation. The premature termination of the trial with significantly fewer patients, might bring a bias into the interpretation. There was also an imbalance in terms of higher age and higher occurrence of pre-stroke disabilities in patients receiving tenecteplase.^16^ These imbalances have been partly compensated by merging the subjects from both trials, achieving better balanced demographics along with a larger and more homogenous cohort, thus yielding a higher statistical power.

The mortality at 3 months was higher in the tenecteplase arm in both the ITT and the PP analyses, which corresponds to the result of NOR-TEST 2A. Moreover, both analyses favor the alteplase in terms of favorable outcome using mRS at 3 months expressed by ordinal shift analysis as well as mRS with cut-off point 0-2. After stratification based on NIHSS score, higher mortality was observed only in severe stroke patients (Figure 3). A similar finding was observed in a previous sub-analysis of NOR-TEST.^15^ However, in NOR-TEST, the amount of any ICH and sICH was more balanced between the arms and sICH was the cause of death only in one patient treated with tenecteplase. The reason for the difference between the trials is not clear. We could not prove that high age has a consequence for the safety of the tenecteplase 0.4mg/kg (Figure 2). This corresponds to the previous studies of alteplase in the elderly, and also to a previous sub-analysis of NOR-TEST.^23, 24^ In the pooled analysis, any ICH and sICH were more common in the middle age group (Figure 2). This group’s predominance in the trials gives statistically stronger results and thus overall negative safety profile of tenecteplase 0.4 mg/kg may be present independently of the age. A by chance observation may, however, also explain the results, since the stratified cohorts, predominantly those ≤60 and ≥80 years, may be underpowered.

There was no significant change in primary and secondary outcome after exclusion of patients with unknown onset of stroke, indicating that these two populations may be similar regarding primary and secondary outcomes. Patients with unknown onset of stroke, however, represent quite small portion and it is therefore not possible to draw any firm conclusion based on the results (Table S1).

Even though there was a higher number of both any ICH and sICH in the tenecteplase arm, sICH was considered as direct cause of death in 4 patients treated with tenecteplase and in 5 patients treated with alteplase. Many patients in the tenecteplase arm died as a consequence of an initial large ischemic stroke or malignant edema, but the cause of death was not attributed to tenecteplase as such.

The frequency of sICH was similar between the pooled analysis and the 0.4 mg/kg tenecteplase arm in EXTEND-IA TNK trial part 2.^25^ However, these two studies are difficult to compare because only 20% of patients in the pooled analysis underwent EVT and a high proportion of these patients had moderate stroke with lower NIHSS on admission. Furthermore, the NOR-TEST trials contain some patients with large vessel occlusion not undergoing EVT for various reasons (Table 1). One can hypothesize that some patients not receiving EVT, mostly having smaller clots, may have delayed recanalization and increased blood brain barrier damage and thereby have higher bleeding rates when tenecteplase is given alone, and the dose 0.4 mg/kg may be too high. This higher bleeding rate might be comparable to the bleeding rate with EVT plus tenecteplase in patients with larger clots. In the NOR-TEST trials, patients undergoing EVT and receiving alteplase more often achieved major neurological improvement and had lower amount of any ICH (Table S2). There was, however, no significant difference in sICH and mortality between the arms. Interestingly, the mortality was similar in patients undergoing EVT and receiving tenectaplese 0.4 mg/kg compared to the same treatment arm in EXTEND-IA TNK trial part 2 (17.8% vs 17%), however the occurrence of sICH was higher in the pooled analysis (8.9% vs 4.7%). The population size difference may be the reason for the difference, as our finding may be underpowered.

The risk of sICH increases with stroke severity, but the treatment benefit of alteplase still outweighs the risk of adverse events independently of age.^26, 27^ Our findings emphasize that stroke severity plays a crucial role when it comes to safety of tenecteplase. In the pooled analysis, any ICH, sICH and mortality at 90 days appeared to be more common in patients with severe stroke when treated with tenecteplase. Neither NOR-TEST 2A nor the pooled analysis therefore could prove that tenecteplase 0.4 mg/kg is non-inferior to standard-dose alteplase regarding safety. Based on these results, we cannot recommend further trials testing the tenecteplase 0.4 mg/kg in acute ischemic stroke.

Patients with mild stroke or stroke mimics treated with alteplase have low occurrence of unfavorable outcome and ICH.^22, 28^ But even though tenecteplase 0.4 mg/kg and standard dose alteplase have similar safety profiles in these populations^13, 21^, further testing of 0.4 mg/kg in these patients also does not seem justifiable.

The convenience of tenecteplase in clinical practice, and its pharmacological superiority, makes tenecteplase a desirable thrombolytic drug in acute stroke therapy.^3^ A lower tenecteplase dose may have a better safety profile but might also have lower efficacy. The ENCHANTED Study did not show non-inferiority of low-dose alteplase to standard-dose alteplase for death and disability at 90 days, but showed significantly fewer symptomatic intracerebral hemorrhages with low-dose alteplase.^29^ However, in a general stroke population, the recently published AcT trial testing tenecteplase 0.25 mg/kg compared to standard-dose alteplase showed non-inferiority in terms efficacy but a positive shift in the safety profile.^30^ In patients with large vessel occlusion treated with intravenous tenecteplase before EVT, the EXTEND-IA TNK part 2 trial suggests that tenecteplase 0.40-mg/kg does not confer an advantage over the 0.25-mg/kg dose.^25^ Thus, tenecteplase 0·25 mg/kg seems to be a reasonable alternative to alteplase for all patients presenting with acute ischaemic stroke and meeting standard criteria for thrombolysis.

In conclusion, the pooled analysis of NOR-TEST and NOR-TEST 2A, indicate a worse safety profile of tenecteplase 0.4 mg/kg as compared to standard dose alteplase in acute ischemic stroke within 4.5 hours after stroke onset, and predominantly so in patients with severe stroke.

## Data Availability

Anonymized data supporting our findings in the presented article may be provided by Vojtech Novotny or Christopher Elnan Kvistad upon reasonable request.

## Sources of funding

The Norwegian National Programme for Clinical Therapy Research (grant reference KLINBEFORSK)

## Disclosures

V. Novotny, C.E. Kvistad, A. Fromm. and H.Næss report no disclosures relevant to the manuscript. L.Thomassen reports an unrestricted grant from Boehringer Ingelheim.

## Notes

### Competing Interest Statement

The authors have declared no competing interest.

### Clinical Trial

NCT01949948, NCT03854500

### Author Declarations

The study was approved by the regional Committee for Medical and Health Research Ethics and the Norwegian Medicines Agency.

## References

1. Hill MD, Buchan AM, Canadian Alteplase for Stroke Effectiveness Study I. Thrombolysis for acute ischemic stroke: Results of the canadian alteplase for stroke effectiveness study. CMAJ. 2005;172:1307–1312

2. Wahlgren N, Ahmed N, Davalos A, Ford GA, Grond M, Hacke W, et al. Thrombolysis with alteplase for acute ischaemic stroke in the safe implementation of thrombolysis in stroke-monitoring study (sits-most): An observational study. Lancet. 2007;369:275–282

3. Logallo N, Kvistad CE, Thomassen L. Therapeutic potential of tenecteplase in the management of acute ischemic stroke. CNS Drugs. 2015;29:811–818

4. Tanswell P, Modi N, Combs D, Danays T. Pharmacokinetics and pharmacodynamics of tenecteplase in fibrinolytic therapy of acute myocardial infarction. Clin Pharmacokinet. 2002;41:1229–1245

5. Haley EC, Jr., Lyden PD, Johnston KC, Hemmen TM, Investigators TNKiS. A pilot dose-escalation safety study of tenecteplase in acute ischemic stroke. Stroke. 2005;36:607–612

6. Haley EC, Jr., Thompson JL, Grotta JC, Lyden PD, Hemmen TG, Brown DL, et al. Phase iib/iii trial of tenecteplase in acute ischemic stroke: Results of a prematurely terminated randomized clinical trial. Stroke. 2010;41:707–711

7. Coutts SB, Dubuc V, Mandzia J, Kenney C, Demchuk AM, Smith EE, et al. Tenecteplase-tissue-type plasminogen activator evaluation for minor ischemic stroke with proven occlusion. Stroke. 2015;46:769–774

8. Parsons M, Spratt N, Bivard A, Campbell B, Chung K, Miteff F, et al. A randomized trial of tenecteplase versus alteplase for acute ischemic stroke. N Engl J Med. 2012;366:1099–1107

9. Huang X, Cheripelli BK, Lloyd SM, Kalladka D, Moreton FC, Siddiqui A, et al. Alteplase versus tenecteplase for thrombolysis after ischaemic stroke (attest): A phase 2, randomised, open-label, blinded endpoint study. Lancet Neurol. 2015;14:368–376

10. Campbell BCV, Mitchell PJ, Churilov L, Yassi N, Kleinig TJ, Dowling RJ, et al. Tenecteplase versus alteplase before thrombectomy for ischemic stroke. N Engl J Med. 2018;378:1573–1582

11. Seners P, Caroff J, Chausson N, Turc G, Denier C, Piotin M, et al. Recanalization before thrombectomy in tenecteplase vs. Alteplase-treated drip-and-ship patients. J Stroke. 2019;21:105–107

12. Burgos AM, Saver JL. Evidence that tenecteplase is noninferior to alteplase for acute ischemic stroke: Meta-analysis of 5 randomized trials. Stroke. 2019;50:2156–2162

13. Logallo N, Novotny V, Assmus J, Kvistad CE, Alteheld L, Ronning OM, et al. Tenecteplase versus alteplase for management of acute ischaemic stroke (nor-test): A phase 3, randomised, open-label, blinded endpoint trial. Lancet Neurol. 2017;16:781–788

14. Anderson CS. Nor-test-ing tenecteplase in acute ischaemic stroke. Lancet Neurol. 2017;16:762–763

15. Kvistad CE, Novotny V, Kurz MW, Ronning OM, Thommessen B, Carlsson M, et al. Safety and outcomes of tenecteplase in moderate and severe ischemic stroke. Stroke. 2019;50:1279–1281

16. Kvistad CE, Naess H, Helleberg BH, Idicula T, Hagberg G, Nordby LM, et al. Tenecteplase versus alteplase for the management of acute ischaemic stroke in norway (nor-test 2, part a): A phase 3, randomised, open-label, blinded endpoint, non-inferiority trial. Lancet Neurol. 2022;21:511–519

17. Logallo N, Kvistad CE, Nacu A, Naess H, Waje-Andreassen U, Asmuss J, et al. The norwegian tenecteplase stroke trial (nor-test): Randomised controlled trial of tenecteplase vs. Alteplase in acute ischaemic stroke. BMC Neurol. 2014;14:106

18. Davalos A, Toni D, Iweins F, Lesaffre E, Bastianello S, Castillo J. Neurological deterioration in acute ischemic stroke: Potential predictors and associated factors in the european cooperative acute stroke study (ecass) i. Stroke. 1999;30:2631–2636

19. Boysen G, Group ES. European cooperative acute stroke study (ecass): (rt-pa-thrombolysis in acute stroke) study design and progress report. Eur J Neurol. 1995;1:213–219

20. Bluhmki E, Chamorro A, Davalos A, Machnig T, Sauce C, Wahlgren N, et al. Stroke treatment with alteplase given 3.0-4.5 h after onset of acute ischaemic stroke (ecass iii): Additional outcomes and subgroup analysis of a randomised controlled trial. Lancet Neurol. 2009;8:1095–1102

21. Kvistad CE, Novotny V, Naess H, Hagberg G, Ihle-Hansen H, Waje-Andreassen U, et al. Safety and predictors of stroke mimics in the norwegian tenecteplase stroke trial (nor-test). Int J Stroke. 2019;14:508–516

22. Logallo N, Kvistad CE, Naess H, Waje-Andreassen U, Thomassen L. Mild stroke: Safety and outcome in patients receiving thrombolysis. Acta Neurol Scand Suppl. 2014:37–40

23. Ford GA, Ahmed N, Azevedo E, Grond M, Larrue V, Lindsberg PJ, et al. Intravenous alteplase for stroke in those older than 80 years old. Stroke. 2010;41:2568–2574

24. Thommessen B, Naess H, Logallo N, Kvistad CE, Waje-Andreassen U, Ihle-Hansen H, et al. Tenecteplase versus alteplase after acute ischemic stroke at high age. Int J Stroke. 2021;16:295–299

25. Campbell BCV, Mitchell PJ, Churilov L, Yassi N, Kleinig TJ, Dowling RJ, et al. Effect of intravenous tenecteplase dose on cerebral reperfusion before thrombectomy in patients with large vessel occlusion ischemic stroke the extend-ia tnk part 2 randomized clinical trial. Jama-J Am Med Assoc. 2020;323:1257–1265

26. Ingall TJ, O’Fallon WM, Asplund K, Goldfrank LR, Hertzberg VS, Louis TA, et al. Findings from the reanalysis of the ninds tissue plasminogen activator for acute ischemic stroke treatment trial. Stroke. 2004;35:2418–2424

27. Emberson J, Lees KR, Lyden P, Blackwell L, Albers G, Bluhmki E, et al. Effect of treatment delay, age, and stroke severity on the effects of intravenous thrombolysis with alteplase for acute ischaemic stroke: A meta-analysis of individual patient data from randomised trials. Lancet. 2014;384:1929–1935

28. Zinkstok SM, Engelter ST, Gensicke H, Lyrer PA, Ringleb PA, Artto V, et al. Safety of thrombolysis in stroke mimics: Results from a multicenter cohort study. Stroke. 2013;44:1080–1084

29. Anderson CS, Robinson T, Lindley RI, Arima H, Lavados PM, Lee TH, et al. Low-dose versus standard-dose intravenous alteplase in acute ischemic stroke. N Engl J Med. 2016;374:2313–2323

30. Menon BK, Buck BH, Singh N, Deschaintre Y, Almekhlafi MA, Coutts SB, et al. Intravenous tenecteplase compared with alteplase for acute ischaemic stroke in canada (act): A pragmatic, multicentre, open-label, registry-linked, randomised, controlled, non-inferiority trial. Lancet. 2022;400:161–169

